# Testing key messages about extending cervical screening intervals

**DOI:** 10.1101/2022.01.12.22269122

**Authors:** Laura Marlow, Martin Nemec, Jessica Barnes, Jo Waller

## Abstract

**Objectives:** We tested the impact of different messages about the rationale for extended screening intervals on acceptability of an extension.

**Methods:** Women in England aged 25-49 years (n=2931) were randomised to read different messages about extending intervals from 3 to 5 years. Outcome measures were general acceptability and six components from the Theoretical Framework of Acceptability (TFA).

**Results:** The control group were less likely to find the change acceptable (43%) than the groups who saw additional messages (47-63%). Women who saw messages about interval safety, test accuracy or the speed of cell changes had more positive affective attitudes, higher ethicality beliefs, a better understanding of the reasons for the interval change and were more likely to believe that 5-year intervals would be safe. Being up-to-date with screening and previous abnormal results were associated with finding 5-yearly screening unacceptable.

**Conclusions:** Emphasising the slow development of cell changes following an HPV negative result and the safety of longer intervals, alongside messages about the accuracy of HPV primary screening is important.

**Practical implications:** Campaigns explaining the rationale for increased screening intervals are likely to improve acceptability. Though some women who feel at increased risk, may remain worried even when the rationale is explained.

## INTRODUCTION

In 2019, England’s cervical screening programme changed to HPV Primary screening (PHE, 2016). The greater sensitivity of HPV primary screening means that extending screening intervals for women aged 25-49 years from 3-yearly to 5-yearly is likely to be safe. Several other countries have already implemented longer intervals including Australia and the Netherlands. Previous work suggests that transitions to longer screening intervals are not always considered acceptable to screening participants and can result in worry and concern (Obermair et al., 2018; Ogilvie et al., 2016; Silver et al., 2015). However, acceptability of extended screening intervals can be improved by explaining the rationale for changes (Hill et al., 2021) and by involving women in the development of communication strategies designed to explain program changes (Rachael H Dodd et al., 2020).

Acceptability “reflects the extent to which people delivering or receiving a healthcare intervention consider it to be appropriate, based on anticipated or experienced cognitive and emotional responses” (Sekhon et al., 2017) and is an important consideration ahead of policy changes. The Theoretical Framework of Acceptability (TFA) conceptualises acceptability as a multi-dimensional construct encompassing seven components: ethicality, affective attitude, burden, intervention coherence, opportunity costs, perceived effectiveness and self-efficacy (Sekhon et al., 2018).

Building on an exploratory qualitative study (Nemec et al., 2021), we developed and tested five prototype messages to inform women about the planned change to cervical screening intervals. The aim of this study was to establish which of these messages were most likely to improve acceptability of the cervical screening interval to help inform communication approaches in the UK and elsewhere.

## METHODS

### Design

A quasi-experimental online study was used, randomising women to read different messages about extended cervical screening intervals. A protocol for the study is available on Open Science Framework (https://osf.io/2p7ba/).

### Participants

Participants were members of a research panel (maintained by Dynata Global UK Ltd) and were invited to participate in an online study via email. Participants clicked on a link and were directed to complete the survey which was hosted on SurveyMonkey. Following consent, we assessed eligibility. Eligible participants self-identified as female; were 25-49 years; living in England; and had no previous diagnosis of cervical cancer. We expected 30-50% to be the lowest possible proportion of women who would find the interval change to be acceptable. With alpha=.05 and power of 85%, we needed approximately 427-463 women per group to detect a 10% difference. We anticipated an exclusion rate of 10%, and commissioned recruitment of 3087 participants, approximately 516 in each exposure group.

### Procedure

Eligible participants completed baseline measures and read two ‘announcement’ messages introducing HPV testing and the planned interval change. Participants were then randomised to one of six groups. The ‘control’ group saw no further messages. The remaining five groups each saw one additional message (see Table 1 for all messages). Participants then responded to questions assessing acceptability of the cervical screening interval change before being presented with all five messages and asked to rank them from 1 (most important) to 5 (least important).

**Table 1:** Messages about HPV and the rationale for extended intervals.

|  |
| --- |
| Control group - Announcement messages only<br>*Cervical screening used to start by looking for abnormal cells in the cervix, but in 2019, the process changed. Nearly all cervical cancers are caused by a virus called human papillomavirus (HPV). The screening test itself is carried out in the same way as before but we now start by checking the sample for the types of HPV that can cause cervical cancer. This means we can make further changes to improve cervical screening for you.*<br>*From 2021, people your age (25-49 years) will be invited for screening every 5 years instead of every 3 years.*<br>Timeline message<br>*There are three steps in the development of cervical cancer. The first is infection with HPV. If your immune system does not get rid of the virus, HPV can cause the second step, abnormal cells. Thirdly, if the abnormal cells are not treated, they can turn into cancer. The development of HPV into cervical cancer is usually a slow process taking over 10 years.*<br>If HPV Positive message<br>*If your sample is HPV positive (i.e. you have HPV), we will also check it for abnormal cervical cells. If none are found, you will be asked to come for screening again in 12-months. This is to check if your immune system has got rid of HPV. If you have HPV and abnormal cervical cells we will refer you for another examination.*<br>Accuracy message<br>*Before HPV testing, screening would occasionally miss abnormal cells, so it was important for you to come for screening every 3 years in case this happened. The HPV test is better at picking up if you have HPV so you do not need to have screening as often.*<br>Safety message<br>*If you have an HPV negative screening result (i.e. you do not have HPV), it is safe to wait five years until your next screen. Even if you have had abnormal cells before, or had treatment, it is still safe for you to have 5 yearly screening if you are HPV negative. You only need further tests if you have HPV.*<br>Speed of cell changes message<br>*HPV progresses very slowly. This means any abnormal cells in the cervix that develop within the 5 years between your screening tests can still be treated when picked up at the next test. Having screening 3 years after a negative HPV result is usually too soon for cells in your cervix to become abnormal.* |

Note: All participants saw the ‘Announcement messages’

### Exposure

Messages were developed following qualitative work (Nemec et al., 2021). This work highlighted key information that women felt they would need to know about extended screening intervals. This included addressing the questions: Why now and not before? Is it safe? How quickly do cell changes occur? Could abnormalities develop between tests and could HPV be ‘missed’? What is the procedure for women with positive results? Messages were developed to address five different themes: Timeline, What happens when you are HPV positive, Accuracy, and Speed of changes. The wording of the messages reflected terminology used in the NHS cervical screening information leaflet and was refined following feedback from women in the eligible age range (n=19) and stake holders working in the cervical screening programme (n=7). Table 1 shows the final messages used.

### Measures

The questionnaire is on OSF (https://osf.io/vn6d3/). A single item assessed overall acceptability at baseline, after exposure and again after seeing all messages: ‘How acceptable do you feel it is to have cervical screening every 5 years?’ (Completely unacceptable; Unacceptable; Acceptable; Completely acceptable; no opinion). Participants were recoded into those who found the 5-year interval ‘acceptable’ versus ‘unacceptable’. ‘No Opinion’ was treated as missing.

An additional 18 items were developed to assess the relevant components of acceptability identified by the Theoretical Framework of Acceptability (TFA). This included *Affective attitude* (how an individual feels about the intervention); *Ethicality* (is the intervention a good fit with the individual’s value system); *Self-efficacy* (is the individual confident they can perform the intervention); *Intervention coherence* (the extent to which an individual understands the intervention), *perceived effectiveness* (is the intervention likely to achieve its purpose) and *opportunity cost* (does the individual need to give up any benefits or values). Items were designed for this study and informed by a generic measure of acceptability (unpublished), a previous study assessing cervical screening interval changes (Hill et al., 2021) and our exploratory qualitative work (Nemec et al., 2021). Responses to each item were on a 4-point scale (strongly disagree to strongly agree, with an addition option for ‘no opinion’). Items were presented in a random order.

Participants completed measures assessing age, education, sexual orientation, relationship status, work status, ethnicity, screening history, screening intentions, awareness of HPV and HPV vaccination status. We also included 4 items assessing knowledge of the change and 3-items assessing understanding of how quickly HPV develops into cervical cancer (timeline).

### Analysis

A full analysis plan was pre-registered on OSF prior to receipt of data (https://osf.io/fnbj9/). Analyses were carried out in SPSS v25.

We used logistic regression to explore whether exposure group (6-levels, with control group as the reference category) was associated with finding the change to screening intervals ‘acceptable’ versus ‘unacceptable’. This was run i) unadjusted, ii) adjusted for age, education, screening history and vaccination status iii) adjusted for age, education, screening history, vaccination status, knowledge of the change and understanding of how quickly cervical cancer develops (timeline).

We also used multiple or logistic regression to look at whether exposure group was associated with each of the six components of acceptability based on the TFA. Cronbach’s alpha was good for affective attitude (0.92), ethicality (0.88), self-efficacy (0.68) and knowledge of the change (0.69) so these were treated as combined continuous scales (range 1-5, created using the mean). All other items are presented as individual categorical variables and were recoded into binary variables (‘agree/strongly agree’ versus ‘disagree/strongly disagree’). We also looked at whether any of the messages were consistently ranked higher or lower than others on importance using the Friedman Test.

We looked at whether there was a significant difference in the proportion of women who would find the change acceptable before and after having seen all of the information using the McNemar Test (among women in the control group only, n=375) and explored whether there were subgroups of women who continued to find the change to 5-yearly screening unacceptable after seeing all of the information (using logistic regression adjusting for initial message exposure).

## RESULTS

### Sample Characteristics

A total of 3440 women started the survey. We excluded those without consent (n=88), those not eligible (n=186) and those who completed the survey too slowly/quickly/did not read the message (n=235). This left data from 2931 for analyses. Mean age was 37.8 years (SD=6.9) and 71% of participants were up to date with cervical screening (i.e. had a test within the last 3 years). Sample characteristics are shown in Table 2.

**Table 2:** **Sample Characteristics (n=2,931) and percentage of women who would find 5-yearly screening unacceptable after seeing all five messages**.

|  | Whole sample<br>n (%) | n (%) who find 5-yearly<br>screening unacceptable<sup>a</sup> | Odds Ratio<br>(95% CI)<sup>b</sup> |
| --- | --- | --- | --- |
| Age group |  |  |  |
| 25-29 years | 427 (14.6) | 121 (30.8) | 1.11 (0.86-1.43) |
| 30-39 years | 1257 (42.9) | 364 (31.6) | 1.13 (0.94-1.35) |
| 40-49 years | 1247 (42.5) | 324 (28.9) | 1.00 |
| Educational level |  |  |  |
| Low-level | 564 (19.2) | 160 (32.9) | 1.25 (1.00-1.57) |
| Mid-level | 987 (33.7) | 286 (31.6) | 1.17 (0.97-1.41) |
| High-level | 1357 (46.3) | 356 (28.3) | 1.00 |
| Sexual Orientation |  |  |  |
| Heterosexual/Straight | 2695 (91.9) | 754 (30.6) | 1.00 |
| Gay, Lesbian, Bisexual, other | 189 (6.4) | 45 (26.8) | 0.83 (0.58-1.18) |
| Marital Status |  |  |  |
| Single | 857 (29.2) | 211 (27.8) | 1.00 |
| Married/civil partnership/cohabiting | 1897 (64.7) | 556 (31.8) | 1.89 (0.98-1.44) |
| Separated/Divorced/Widowed | 158 (5.4) | 41 (27.7) | 1.01 (0.68-1.50) |
| Work Status |  |  |  |
| Employed | 2148 (73.3) | 603 (30.5) | 1.00 |
| Not Working | 736 (25.1) | 195 (29.9) | 0.99 (0.82-1.20) |
| Ethnic background |  |  |  |
| White British | 2197 (75.0) | 618 (30.8) | 1.00 |
| Any other White background | 274 (9.3) | 87 (34.9) | 1.20 (0.91-1.59) |
| Mixed ethnic background | 78 (2.7) | 25 (34.7) | 1.25 (0.76-2.05) |
| Any Asian Background | 216 (7.4) | 39 (19.9) | 0.56 (0.39-0.81) |
| Any Black background | 59 (2.0) | 12 (23.1) | 0.68 (0.35-1.31) |
| Other | 71 (2.4) | 20 (31.8) | 1.00 (0.58-1.72) |
| Screening Status |  |  |  |
| Up to date | 2076 (70.8) | 652 (34.0) | 1.00 |
| Overdue | 450 (15.4) | 88 (21.4) | 0.53 (0.41-0.69) |
| Non-attender | 364 (12.4) | 62 (20.1) | 0.49 (0.36-0.66) |
| Screening intention next time invited |  |  |  |
| Intender | 2597 (88.6) | 767 (32.0) | 1.00 |
| Non-intender | 334 (11.4) | 42 (15.7) | 0.40 (0.28-0.56) |
| Experience of abnormal cells (yes) |  |  |  |
| No/not sure | 2242 (76.5) | 579 (28.0) | 1.00 |
| Yes | 589 (20.1) | 222 (38.9) | 1.66 (1.37-2.02) |
| Heard of HPV (Yes) |  |  |  |
| Yes | 782 (26.7) | 194 (27.1) | 1.00 |
| No | 2090 (71.3) | 615 (31.5) | 1.24 (1.03-1.50) |
| Vaccinated against HPV (yes) |  |  |  |
| No/Don't know | 2573 (87.8) | 714 (30.0) | 1.00 |
| Yes | 291 (9.9) | 92 (33.2) | 0.87 (0.67-1.14) |

<sup>a</sup> unadjusted; <sup>b</sup> adjusted for exposure group

### Acceptability of 5-yearly screening by key message

Overall acceptability of 5-yearly screening was associated with the message women were exposed to. In the control group, where women saw only the two messages announcing the change, 43% found the change ‘acceptable’ or ‘completely acceptable’. This proportion was significantly higher in each of the groups who saw an additional message (47-63%; See Table 3).

**Table 3:**
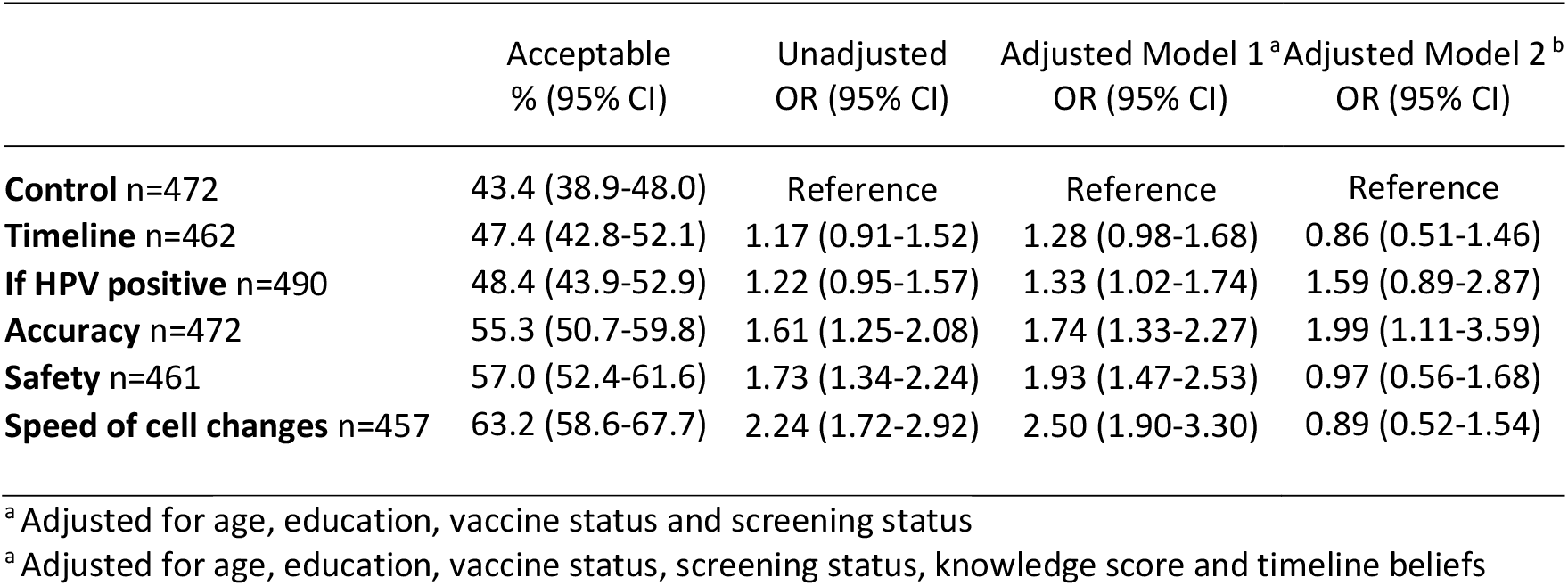
**The association between message exposure and overall acceptability of 5-yearly screening**

|  | Acceptable<br>% (95% CI) | Unadjusted<br>OR (95% CI) | Adjusted Model 1<sup>a</sup><br>OR (95% CI) | Adjusted Model 2<sup>b</sup><br>OR (95% CI) |
| --- | --- | --- | --- | --- |
| **Control** n=472 | 43.4 (38.9-48.0) | Reference | Reference | Reference |
| **Timeline** n=462 | 47.4 (42.8-52.1) | 1.17 (0.91-1.52) | 1.28 (0.98-1.68) | 0.86 (0.51-1.46) |
| **If HPV positive** n=490 | 48.4 (43.9-52.9) | 1.22 (0.95-1.57) | 1.33 (1.02-1.74) | 1.59 (0.89-2.87) |
| **Accuracy** n=472 | 55.3 (50.7-59.8) | 1.61 (1.25-2.08) | 1.74 (1.33-2.27) | 1.99 (1.11-3.59) |
| **Safety** n=461 | 57.0 (52.4-61.6) | 1.73 (1.34-2.24) | 1.93 (1.47-2.53) | 0.97 (0.56-1.68) |
| **Speed of cell changes** n=457 | 63.2 (58.6-67.7) | 2.24 (1.72-2.92) | 2.50 (1.90-3.30) | 0.89 (0.52-1.54) |

<sup>a</sup> Adjusted for age, education, vaccine status and screening status
<sup>b</sup> Adjusted for age, education, vaccine status, screening status, knowledge score and timeline beliefs

There were also statistically significant between-group differences in affective attitudes, ethicality, intervention coherence and perceptions of whether the 5-yearly interval is safe (See Table 4). Women who saw the safety message, the accuracy message or the speed of cell changes message had more positive affective attitudes, higher ethicality beliefs, a better understanding of the reasons for why the interval change was happening and were more likely to believe that 5-yearly intervals would be safe. The timeline message improved understanding of why the changes were being made.

**Table 4:**
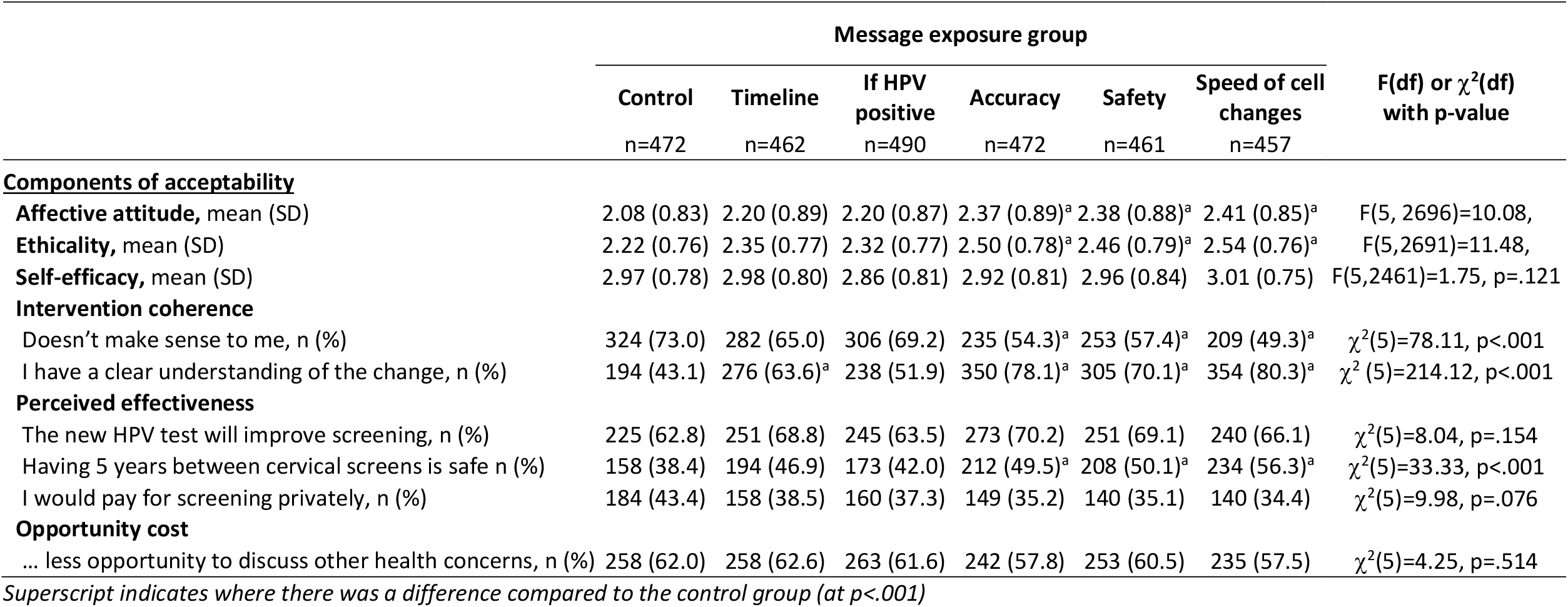
**The impact of five different messages about HPV and the rationale for extended intervals on acceptability of 5-yearly screening in England (unadjusted)**

| | Message exposure group | | | | | | F(df) or $\chi^2$(df) with p-value |
| --- | --- | --- | --- | --- | --- | --- | --- |
|  | Control | Timeline | If HPV positive | Accuracy | Safety | Speed of cell changes |  |
|  | n=472 | n=462 | n=490 | n=472 | n=461 | n=457 |  |
| **Components of acceptability** |  |  |  |  |  |  |  |
| **Affective attitude**, mean (SD) | 2.08 (0.83) | 2.20 (0.89) | 2.20 (0.87) | 2.37 (0.89)<sup>a</sup> | 2.38 (0.88)<sup>a</sup> | 2.41 (0.85)<sup>a</sup> | F(5, 2696)=10.08, |
| **Ethicality**, mean (SD) | 2.22 (0.76) | 2.35 (0.77) | 2.32 (0.77) | 2.50 (0.78)<sup>a</sup> | 2.46 (0.79)<sup>a</sup> | 2.54 (0.76)<sup>a</sup> | F(5,2691)=11.48, |
| **Self-efficacy**, mean (SD) | 2.97 (0.78) | 2.98 (0.80) | 2.86 (0.81) | 2.92 (0.81) | 2.96 (0.84) | 3.01 (0.75) | F(5,2461)=1.75, p=.121 |
| **Intervention coherence** |  |  |  |  |  |  |  |
| Doesn't make sense to me, n (%) | 324 (73.0) | 282 (65.0) | 306 (69.2) | 235 (54.3)<sup>a</sup> | 253 (57.4)<sup>a</sup> | 209 (49.3)<sup>a</sup> | $\chi^2$(5)=78.11, p<.001 |
| I have a clear understanding of the change, n (%) | 194 (43.1) | 276 (63.6)<sup>a</sup> | 238 (51.9) | 350 (78.1)<sup>a</sup> | 305 (70.1)<sup>a</sup> | 354 (80.3)<sup>a</sup> | $\chi^2$ (5)=214.12, p<.001 |
| **Perceived effectiveness** |  |  |  |  |  |  |  |
| The new HPV test will improve screening, n (%) | 225 (62.8) | 251 (68.8) | 245 (63.5) | 273 (70.2) | 251 (69.1) | 240 (66.1) | $\chi^2$(5)=8.04, p=.154 |
| Having 5 years between cervical screens is safe n (%) | 158 (38.4) | 194 (46.9) | 173 (42.0) | 212 (49.5)<sup>a</sup> | 208 (50.1)<sup>a</sup> | 234 (56.3)<sup>a</sup> | $\chi^2$(5)=33.33, p<.001 |
| I would pay for screening privately, n (%) | 184 (43.4) | 158 (38.5) | 160 (37.3) | 149 (35.2) | 140 (35.1) | 140 (34.4) | $\chi^2$(5)=9.98, p=.076 |
| **Opportunity cost** |  |  |  |  |  |  |  |
| ... less opportunity to discuss other health concerns, n (%) | 258 (62.0) | 258 (62.6) | 263 (61.6) | 242 (57.8) | 253 (60.5) | 235 (57.5) | $\chi^2$(5)=4.25, p=.514 |

*Superscript indicates where there was a difference compared to the control group (at p<.001)*

When asked to rank the five messages from most to least important, mean ranks for the messages ranged from 2.91 (SD=1.35) to 3.17 (SD=1.42). The consistency between participants was incredibly low (Kendall’s W=0.01), suggesting there was no overall consensus on the most important message. This was the case overall and when stratified by age, education, screening status or previous experience of abnormal cells.

### Acceptability of 5-yearly screening after reading all five key messages

Among women in the control group, 43% (205/472) said they found 5-yearly screening acceptable after seeing the announcement messages. Once these women had seen all five messages, acceptability was 66% (293/447), representing a statistically significant increase (McNemar Test with n=434, p<.001).

After all women had seen all five messages, 28% (809/2931) still felt that 5-yearly screening was unacceptable. After adjusting for initial exposure group there were no significant associations between demographic characteristics and finding the 5-yearly interval unacceptable (see Table 2). However, participants who were overdue for screening or were non-attenders were less likely to find 5-yearly screening unacceptable (21% and 20% respectively) compared to women who were up-to-date with screening (34%). Women who did not intend to go for screening when next invited were less likely to find 5-yearly screening unacceptable (16%) compared to women who did intend to go when next invited (32%). Women who had past experience with abnormal cells were more likely to find the move to 5-yearly screening unacceptable (39%) compared to women who did not have experience of abnormal cells (28%). Women who had not heard of HPV before were less likely to find the move to 5-yearly screening unacceptable (27%) than women who had heard of HPV before (32%).

## DISCUSSION AND CONCLUSION

### Discussion

Presenting messages designed to explain the rationale for changes to the cervical screening intervals can improve acceptability of the change. Messages about safety, accuracy and speed of cell changes resulted in more positive attitudes and feelings that extended intervals would be safe and ethical, suggesting these are particularly important messages to communicate. No single message was consistently ranked as most important and the highest reported acceptability in the study came after women had seen all five messages.

Adjusting for socio-demographics made very little difference to the impact of message group on acceptability. However, after adjusting for knowledge and timeline, the confidence intervals around overall acceptability percentages widened, and many of the exposure groups were no longer statistically significantly different from the control group. This suggests that the ‘mechanism’ by which each message results in greater acceptability is through improving a greater understanding of the rationale for increased intervals.

After reading all the key messages, around a quarter of women still found a 5-yearly interval unacceptable. This was not statistically associated with age, marital status or socio-economic background. However, women who were up-to-date with screening and intended to continue being screened were less accepting of an extended interval than those who were not regular screening attenders. This is consistent with previous findings (Hill et al., 2021) and suggests reassuring those already engaged in the programme is important e.g. at their appointments and with their results letters.

Nearly 40% of women with previous experience of abnormal cell changes did not consider 5-yearly screening to be acceptable, even after reading all five messages. Despite our attempt to address this concern in one of our messages, explicitly saying: “e*ven if you have had abnormal cells before, or had treatment, it is still safe for you to have 5 yearly screening if you are HPV negative*”. This is consistent with previous work, suggesting that experience of cervical abnormalities can have long lasting effects on women’s perceptions of vulnerability to cervical cancer (Marlow et al., 2021; Obermair et al., 2020; Sharp et al., 2015).

We assessed multiple constructs of acceptability proposed as part of the TFA (Sekhon et al., 2017). This allows us to provide a more holistic picture and is a strength of the study. There are also several limitations of this study. While the online panel recruitment procedure allowed us to randomise message exposure in a controlled way, this does mean our sample is unlikely to be representative of all women aged 25-49 in the English population. However, the sample was diverse, with 25% from ethnic minority backgrounds and 20% having a low level of education. Our previous work suggested that many women were not aware of the move to HPV primary screening and that introducing this concept was an important first step prior to describing a move to 5-yearly screening. For this reason, we provided all participants, including those in the control group, with details of the move to primary screening. It is possible that acceptability would have been different if no additional information had been provided at all. In addition, this work assesses *prospective* acceptability, with the change presented as something that was likely to happen in the future. For most participants, this will have been their first exposure to the idea of a potential increase in interval length. It is possible that once the change is introduced, acceptability will be higher as women grow used to the idea and it becomes normalised.

## Conclusion

In conclusion, this study suggests that presenting messages to address concerns about safety, accuracy and the speed that abnormal changes can develop after acquiring HPV will support acceptability of an extended cervical screening interval. These suggestions could be incorporated into public health messages designed to explain the changes when these are implemented in the future. Subgroups of women defined by their screening history, experiences and intentions are likely to need additional reassurances that increased intervals are safe.

## Practical implications

The findings suggest that the optimal approach to communication about the policy change needs a multi-message campaign. Greater personalisation of information about the interval change, or discussion with a health care professional may be needed to provide reassurance for women who have previously experienced abnormal cells changes. Training to ensure health care professionals are confident in explaining the rationale for extended intervals is important (R H. Dodd et al., 2020).

## Data Availability

The Data and syntax that correspond with the analyses reported will be available on Open Science
Framework following peer review.

## ACKNOWLEDGEMENTS

We would like to thank the PPI representatives and stake holders who took the time to contribute to the design of this study.

## DATA STATEMENT

The Data and syntax that correspond with the analyses reported here are available Open Science Framework.

